# Real-World Performance of Large Language Models in Emergency Department Chest Pain Triage

**DOI:** 10.1101/2024.04.24.24306264

**Authors:** Xiangbin Meng, Jia-ming Ji, Xiangyu Yan, Hua Xu, Jun gao, Junhong Wang, Jingjia Wang, Xuliang Wang, Yuan-geng-shuo Wang, Wenyao Wang, Jing Chen, Kuo Zhang, Da Liu, Zifeng Qiu, Muzi Li, Chunli Shao, Yaodong Yang, Yi-Da Tang

**Affiliations:** Department of Cardiology and Institute of Vascular Medicine, Peking University Third Hospital; State Key Laboratory of Vascular Homeostasis and Remodeling, Peking University; Institute for Artificial Intelligence, Peking University; Institute of Disaster and Emergency Medicine, Tianjin University; Emergency department, Peking university third hospital; Department of Cardiology, State Key Laboratory of Cardiovascular Disease, Fuwai Hospital, National Center for Cardiovascular Diseases, Chinese Academy of Medical Sciences and Peking Union Medical College; Department of Cardiology, the First Hospital of Hebei Medicical University, Graduate School of Hebei Medical University; Peking University Health Science Center, Peking University First Hospital; Peking University Health Science Center, Peking University People’s Hospital

**Author notes:** ***Corresponding to:* Yi-Da Tang, MD, PhD,** Department of Cardiology and Institute of Vascular Medicine, Peking University Third Hospital. State Key Laboratory of Vascular Homeostasis and Remodeling, Peking University, Beijing, China. No.49 Huayuanbei Road, Beijing 100191, China. **Yao-dong Yang, PhD,** Institute for Artificial Intelligence, Peking University, Beijing, China. No.5 Yi HeYuan Road, Beijing 100871, China. These authors contributed equally to this study (Xiangbin Meng, Jia-ming Ji, Xiangyu Yan).

**Keywords:** Large Language Models (LLMs), Real-World Performance, Emergency Department Chest Pain Triage, Acute Coronary Syndrome (ACS), Diagnostic capabilities

## Abstract

**Background:** Large Language Models (LLMs) are increasingly being explored for medical applications, particularly in emergency triage where rapid and accurate decision-making is crucial. This study evaluates the diagnostic performance of two prominent Chinese LLMs, “Tongyi Qianwen” and “Lingyi Zhihui,” alongside a newly developed model, MediGuide-14B, comparing their effectiveness with human medical experts in emergency chest pain triage.

**Methods:** Conducted at Peking University Third Hospital’s emergency centers from June 2021 to May 2023, this retrospective study involved 11,428 patients with chest pain symptoms. Data were extracted from electronic medical records, excluding diagnostic test results, and used to assess the models and human experts in a double-blind setup. The models’ performances were evaluated based on their accuracy, sensitivity, and specificity in diagnosing Acute Coronary Syndrome (ACS).

**Findings:** “Lingyi Zhihui” demonstrated a diagnostic accuracy of 76.40%, sensitivity of 90.99%, and specificity of 70.15%. “Tongyi Qianwen” showed an accuracy of 61.11%, sensitivity of 91.67%, and specificity of 47.95%. MediGuide-14B outperformed these models with an accuracy of 84.52%, showcasing high sensitivity and commendable specificity. Human experts achieved higher accuracy (86.37%) and specificity (89.26%) but lower sensitivity compared to the LLMs. The study also highlighted the potential of LLMs to provide rapid triage decisions, significantly faster than human experts, though with varying degrees of reliability and completeness in their recommendations.

**Interpretation:** The study confirms the potential of LLMs in enhancing emergency medical diagnostics, particularly in settings with limited resources. MediGuide-14B, with its tailored training for medical applications, demonstrates considerable promise for clinical integration. However, the variability in performance underscores the need for further fine-tuning and contextual adaptation to improve reliability and efficacy in medical applications. Future research should focus on optimizing LLMs for specific medical tasks and integrating them with conventional medical systems to leverage their full potential in real-world settings.

## Introduction

Large Language Models (LLMs) are revolutionizing the medical field, particularly in accelerating pre-hospital triage^1–10^. These models leverage deep learning and natural language processing technologies to capture patterns and relationships in text through multi-layer neural network architectures, enabling efficient processing and precise understanding of vast medical data^4–11^. Trained on extensive text data, LLMs have acquired a wealth of vocabulary, grammar, semantics, and a condensed vast knowledge system, allowing them to respond coherently and accurately to various symptom descriptions, medical history information, and literature queries provided by users ^12–15^. Importantly, as the model parameters and training data volume increase, scientists have observed significant “emergence abilities” in LLMs^16–18^. This ability is not only reflected in the efficient processing of complex information but also in their superior performance in logical reasoning and innovative thinking^19^. This gives LLMs unique advantages in simulating human thought patterns, understanding, and applying knowledge, especially in the field of medical diagnostic assistance^20,21^.

Globally, especially in remote areas of developing and developed countries, there is a severe shortage of primary healthcare resources, characterized by insufficient facilities, lack of professional personnel, and financial constraints^22,23^. Trained medical providers, including doctors, nurses, and other community health workers, are scarce, making it difficult to provide high-quality primary healthcare services, further exacerbating the imbalance of medical human resources between urban and rural areas^24^. Traditional clinical prediction models based on machine learning or deep learning, despite having theoretical application prospects, are rarely deployed in actual clinical practice. The main reason is that these models generally lack generalizability and cannot effectively handle the complex and variable actual clinical data, and the required parameters are often not easily obtained in clinical settings^25^. In contrast, LLMs, with their strong information integration and logical reasoning abilities, extensive knowledge reserves, and seamless integration with human language, can overcome these challenges.

In medical diagnostic assistance scenarios, the value of LLMs is particularly prominent. Traditional diagnostic models often struggle to cope with patients’ complex symptom expressions, intricate medical histories, and vast medical literature due to limited processing capacity or insufficient knowledge coverage. LLMs, with their vast training data and deep learning architectures, can quickly organize and integrate various clinical information, using their embedded extensive medical knowledge base to accurately classify and analyze diseases^8,9^. Furthermore, LLMs’ logical reasoning ability allows them to conduct in-depth analysis of complex disease clues, construct disease progression path models, predict potential complications, and provide doctors with detailed and in-depth diagnostic support.

However, a cautiously optimistic attitude should be maintained towards the application of LLMs in the medical environment, fully recognizing their limitations. These limitations include but are not limited to: potential bias in training data, which may lead to unfairness in model decision-making; challenges in explaining complex medical details, which may affect the understanding and trust of doctors and patients in model outputs; and the risk of misdiagnosis due to over-reliance on technology without necessary human supervision. Therefore, while affirming the transformative potential of LLMs in healthcare, it is also crucial to focus on and address these challenges to ensure their application in medical diagnostics is both safe and effective.

A noteworthy challenge arises when applying LLMs to languages such as Chinese, Japanese, Korean, Tamil, Hindi, Thai, and Vietnamese, which employ “non-segmented text” structures that markedly differ from English in terms of grammar, syntax, and usage. Most research endeavors have predominantly concentrated on English-centric models like ChatGPT, leaving a notable research gap in evaluating the diagnostic proficiencies of large language models trained specifically for non-English environments^26,27^. Although English and ‘non-segmented text’ languages share similar fundamental principles in the development of LLMs, they face distinct technical and engineering challenges in practical applications due to differences in language characteristics and available resources. This leads to variations in their implementation and performance. We must consider the fundamental differences in grammatical structures, data resources, vocabulary size and distribution, algorithmic implementation and optimization, as well as the adaptability of technical architectures across different languages. This research gap impacts the broad adoption of these models in diverse linguistic contexts and profoundly influences the level of trust vested in LLMs.

The emergency medical setting is distinguished by its immediacy, intricate clinical presentations, and the imperative need for prompt diagnosis^28^. The environment of emergency room is often dynamic, extremely busy, and high-pressure, requiring healthcare personnel to make rapid decisions and handle multiple cases simultaneously^29^. A study in 2019 found that the average wait time for emergency department patients was approximately 40 minutes before being seen by a physician, with doctors spending an average of 13-24 minutes per patient during the consultation ^30–32^. Emergency triage systems are used globally to assess patient severity and allocate resources^33–35^. The US uses Emergency Severity Index (ESI), a 5-tier system. China uses Emergency Triage Scale/Standard (ETS), a 4-tier system. ETS is like ESI, with levels 1&2 triaged to resuscitation. Patients in levels 3&4 wait to see a physician. Though ETS is generally accurate, some critical patients wait hours and misdiagnosis is a pronounced concern, as research underscores a notably elevated misdiagnosis rate within the emergency room ^36,37^. This issue is exacerbated, particularly for common symptoms associated with myocardial ischemia, which are susceptible to oversight or misjudgment ^38^. The guidelines recommend reperfusion therapy within 12 hours of the onset of myocardial infarction^39,40^, yet approximately 70% of acute myocardial infarction patients succumb to the disease due to the missed opportunity for timely treatment ^41^, highlighting the risk of misdiagnoses leading to treatment delays. LLMs stand poised to bring about significant transformations in specific medical contexts, notably expediting pre-hospital triage procedures. We see potential in these models to facilitate rapid triage by assisting healthcare providers in swiftly processing patient data and offering potential diagnoses rooted in symptoms, medical histories, and pertinent literature.

The medical profession demands precise and dependable tools for informed decision-making. While LLMs hold potential, they present difficulties in understanding context and obtaining clarifications^42–47^. Addressing real-world medical issues requires handling multiple data modalities and must also provide authenticity, authority, accessibility, safety, empathy, and a human touch^48^. Real-world medical problems often transcend the confines of multiple-choice tests and structured tasks. The human or AI model approach to diagnostic interaction, whether single or multi-turn dialogues and their ability to process various data modalities are equally important. Indeed, evaluating these vast medical models might not be any simpler than developing them.

Standardized testing has largely evaluated these models’ medical knowledge reserves and diagnostic logical reasoning capabilities^2,6,8,49^. However, medical issues in the real world often surpass the scope of structured tasks and multiple-choice tests, exhibiting greater complexity and uncertainty^7,50^. Currently, there is a particular lack of systematic evaluation globally on the effectiveness of large language models in real medical environments, especially based on rich, diverse, and dynamically changing real medical data^51^.

In this study, we focused on evaluating two prominent Chinese language models, “Tongyi Qianwen (通•千•) (V1.0.3)“and “Lingyi Zhihui (灵医智慧) (V2.2.0)“^52–54^, which are developed based on a Transformer’s autoregression framework, akin to other globally recognized models like Meta’s LLaMa Series, Google’s PaLM and LaMDA, and OpenAI’s ChatGPT series. As general-purpose large models, they have not been specifically fine-tuned for the medical domain and are currently offered as online services by their respective operators. One of the core objectives of our research is to conduct a comprehensive evaluation of these two models using case data from Chinese patients, focusing particularly on their performance in processing Chinese medical contexts, given that they are primarily trained on Chinese datasets, although they also incorporate a certain proportion of English data. A key component of the study is a comparative analysis of the diagnostic accuracy of “Tongyi Qianwen” and “Lingyi Zhizhi” in handling complex and urgent medical data, benchmarking their results against medical experts’ judgments. Targeted enhancements and optimizations were applied to the fine-tuning and alignment phases, particularly for LLMs tasked with medical applications. Recognizing the performance variability of LLMs underscores the importance of meticulously establishing benchmarks that are apt for medical artificial intelligence. We further integrated the insights gained from the benchmark conducted into developing a new model called **MediGuide-14B**.

## Methods

### Study Design and Setting

This retrospective study was conducted at the Emergency Chest Pain Centers of the Peking University Third Hospital Group, involving five tertiary-level centers. The study received ethical approval from the ethics committee of Peking University Third Hospital (M2023828), complying with the Helsinki Declaration.

### Data Sources

Chief complaints, current medical history, past medical history, family history, and personal history were extracted from electronic medical records as unstructured text content. It is important to note that diagnostic test results such as electrocardiograms, myocardial enzyme tests, and echocardiograms were not included in the test dataset provided to the test model and the control group. Confirmed diagnostic information and related evidence were only made available during the subsequent double-blind evaluation phase to the expert review committee, which served as the authoritative reference standard. This approach aimed to ensure that experts could conduct accurate and fair comparative analyses of the diagnostic accuracy of each group by combining comprehensive and detailed medical test data with the model’s predictive results.

### Participants

The inclusion criteria for this study were patients who visited the emergency chest pain center at Peking University Third Hospital’s five tertiary-level centers due to chest pain-related symptoms between June 2021 and May 2023. The exclusion criteria were: 1.Cases with significant omissions or incompleteness in medical records, such as missing key clinical assessment records or essential auxiliary examination results, which are crucial for a comprehensive patient evaluation; 2. Cases where the chief complaint information did not come directly from the patients themselves due to unstable vital signs or other reasons but was obtained through the accounts of others, making it difficult to accurately trace and complete. Given that such situations could affect the credibility of the study results due to the one-sidedness of the information or decreased accuracy of symptom description, these cases were excluded in the analysis process. Ultimately, 11,428 patients who met the above inclusion and exclusion criteria were included in the study.

### Outcome

The study employed repeated random sampling for case selection, using Python 3.8 to randomly select 100 patient cases from the database, repeated 1000 times. This method ensured a diverse and random sample, reducing potential bias. All medical records were anonymized to maintain patient privacy, further minimizing selection bias and enhancing the generalizability of the findings.

The primary outcome of this study was the accuracy of diagnosing Acute Coronary Syndrome (ACS). The study used LLMs prompts that included patient demographics, clinical symptoms, and medical history, which are commonly found and critically important in primary healthcare and emergency scenarios. This approach mimics the situations where lab results are not yet available, and doctors and medical professionals must rely solely on the patient’s chief complaints and past medical history to triage chest pain and provide rapid management. In the study, the cardiovascular physicians’ group also followed the same information dimensions and problem structure as the LLMs for case analysis. These cardiology specialists, during the diagnostic process, similarly needed to interpret each case based on the patient’s age, gender, chief complaint, present illness history, family history, and personal history, and make diagnostic and therapeutic decisions accordingly.

### Study Size

To ensure a statistical power of 0.8 and effectively compare the diagnostic accuracy between human medical experts and Large Language Models (LLMs) optimized for the medical environment, it is necessary to determine an appropriate sample size. Based on preliminary data and relevant literature, the diagnostic accuracy of human experts usually falls within the range of 0.9 to 0.95, while the accuracy of general-purpose LLMs not specifically trained is between 0.7 and 0.8. LLMs that have been optimized for the medical field have improved their accuracy to levels comparable to human experts, although there may still be slight differences between the two^55^. In this context, to ensure a statistical power of 0.8, sufficient to detect the potential small difference in accuracy between human experts and optimized LLMs, we calculated that each group needs approximately 3835 samples. This sample size ensures that even minor differences in accuracy can be detected with an 80% probability in statistical tests.

Considering the study subjects, we selected five centers affiliated with Peking University Third Hospital, each of which receives about 5000 patients with chest pain annually. Given that clinical data often have high noise, sparsity, and heterogeneity, which not only increase the difficulty of data analysis but also may affect the robustness of the research conclusions, we decided to set the study period from June 2021 to May 2023, totaling two years. This time frame was chosen to accumulate sufficient case data, overcome the inherent complexity of clinical data, and ensure the effectiveness and reliability of the final statistical analysis.

Figure 1 illustrates the flow chart of the entire study. The sample size of 11,428 records was determined based on the hospital’s patient flow and data availability during the study period. This size was deemed sufficient to provide a robust analysis of the LLMs’ diagnostic performance.

**Fig. 1:**
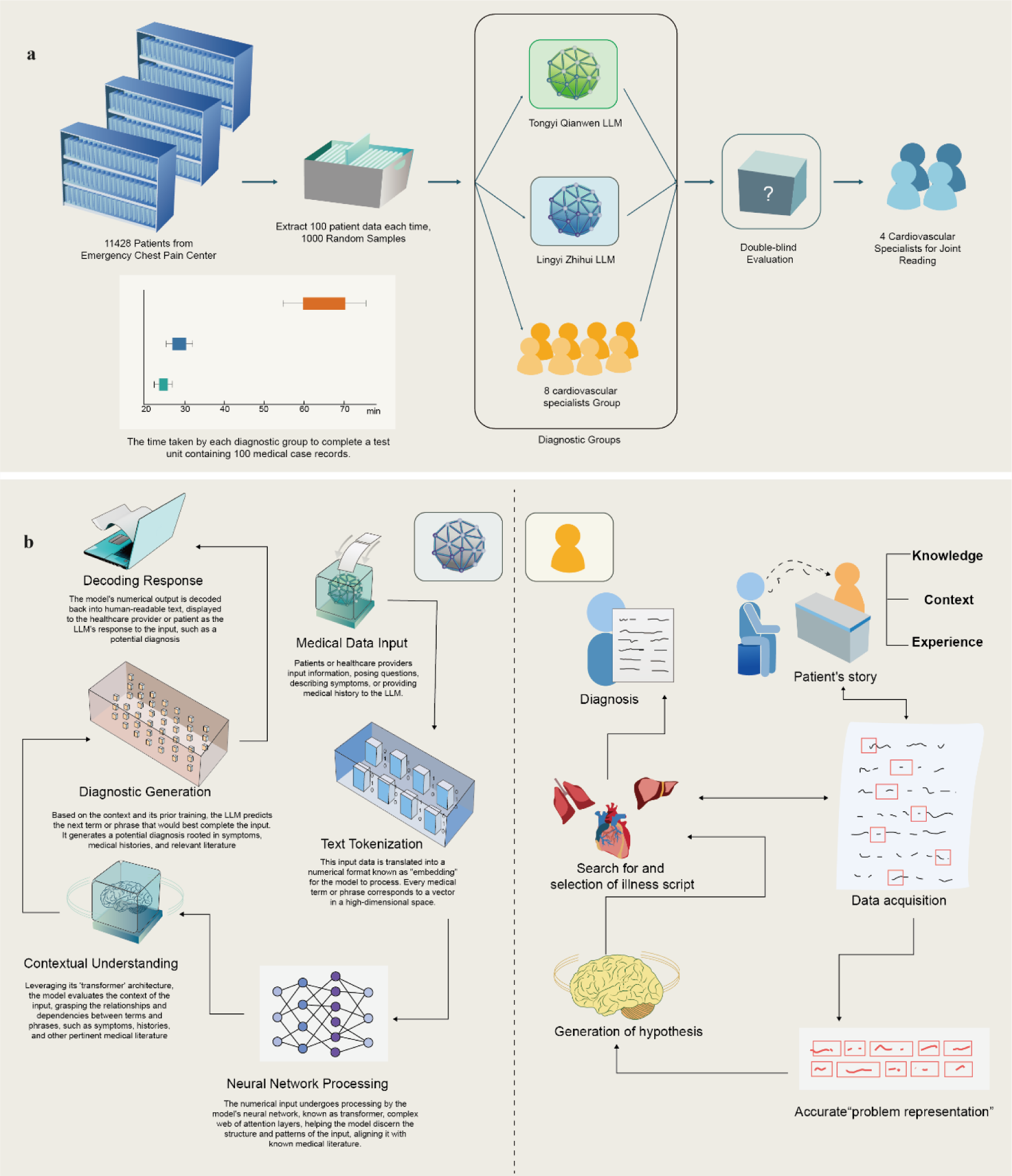
Research process diagram accompanied by a comparative illustration of diagnosis time between LLMs and human experts, along with a comparison of the diagnostic thought processes of LLMs and humans. The research process diagram displays the primary steps and methodologies of this study. The comparative illustration showcases the time disparities between LLMs and human experts in completing diagnostic tasks. The thought process comparison further elucidates the cognitive and decision-making pathways employed by both during diagnosis.

#### LLMs

“Tongyi Qianwen,” developed by Alibaba Group, is a 100 billion-parameter model with a diverse data foundation, including web texts and specialized literature, and utilizes advanced reinforcement learning techniques like A2C/A3C and PPO, and Q-learning methods such as DQN and C51^56,57^.

“Lingyi Zhihui,” created by Baidu Group for medical contexts, has 260 billion parameters. It combines auto-regressive and auto-encoding frameworks, suited for natural language understanding and generation, and supports zero-shot, few-shot learning, and detailed fine-tuning^58,59^.

#### Diagnostic performance

The anonymized dataset, excluding lab test results, was analyzed by “Tongyi Qianwen,” “Lingyi Zhihui,” and a group of human experts. The human experts were eight cardiovascular specialists with over ten years of experience, certified by the Chinese Society of Cardiology and the Chinese Medical Association. Both LLMs and human experts were given detailed patient information encompassing age, gender, chief complaints, present illness, and medical history. In this study, all participants underwent comprehensive training before commencement to ensure a thorough understanding of the research process. During the actual testing phase, we observed and recorded the time taken by each diagnostic group to complete a test unit containing 100 medical case records.

#### Prompt Engineering

For large language models, appropriate prompts are necessary to activate their respective capabilities. (Supplementary Table S1)

The prompt is: “1. Based on the patient’s basic information, chief complaint, symptoms, and medical history, what do you consider to be the patient’s diagnosis? 2. Considering your considered diagnosis, what further tests and medical advice would you recommend? 3. Please evaluate the risk of the patient’s condition based on the above patient information”.

Similarly, a group of human experts will respond to the same questions based on the patient’s basic information, chief complaint, symptoms, and medical history.

#### Reference standards

The gold standard of final diagnosis was the consensus of an independent cardiology expert panel with 20 years of clinical service experience following Chest Pain Management Guidelines^38,60,61^. This panel had full access to patient records and cardiovascular lab test results, ensuring a comprehensive and authoritative standard for comparison. The definitive diagnosis for cases where there was uncertainty was established using Voting Mechanisms. (**Figure 1**)

#### Statistical analysis

The statistical analysis in this study was conducted using the SPSS 27.0 statistical package for Windows, Python version 3.8, and R software version 4.2.2, provided by the R Foundation for Statistical Computing. All continuous variables that adhered to a normal distribution were represented as means with their 95% confidence intervals (CI). To identify initial differences in baseline characteristics between treatment groups, bivariate analyses were performed utilizing Student’s t-test. For comparative analyses among multiple groups, a one-way ANOVA test was employed. A p-value of less than 0.05 was designated as the threshold for statistical significance.

In assessing the differences in diagnostic efficacy for cardiovascular diseases among the study groups, this research employed a comprehensive and multifaceted set of evaluation metrics, including Accuracy, Sensitivity, Specificity, False Positive Rate (FPR), Recall, and confusion matrix diagrams among other multidimensional indicators. These multidimensional indicators together form a rigorous and comprehensive performance evaluation framework, aimed at comprehensively comparing and assessing the strengths and weaknesses of each study group in terms of diagnostic accuracy and effectiveness.

Before evaluating the diagnostic metrics of each group, the study initially assessed the distribution characteristics of the results from 1000 test units in each group using probability density curves, P-P plots, and Q-Q plots to conduct normality tests.

### The Development of MediGuide-14B

MediGuide-14B is developed on the foundation of the Qwen-14B model, undergoing extensive optimization and specialized transformation through a meticulously crafted medical data training and tuning program. Qwen-14B boasts 14 billion model parameters, endowing it with powerful learning capabilities, ample knowledge reserves, and robust logical reasoning. During its foundational training, Qwen-14B assimilated knowledge from over three trillion tokens, spanning Chinese, English, and various other languages, including specialized domains like programming and mathematics. Qwen-14B excels in natural language understanding, mathematical problem-solving, logical reasoning, and computer programming. This base model supports comprehensive fine-tuning, allowing for deep and customized adjustments tailored to various tasks and domains.

The special medical database built by the research team includes detailed medical records of 105,290 outpatients and inpatients, totaling 2 million pieces of professional medical data that have been carefully cleaned and protected for privacy. The training of MediGuide-14B is completed on a high-performance server equipped with A800 80G*8. Leveraging the power of the DeepSpeed framework and the Transformer architecture, we have optimized MediGuide-14B for better performance and efficiency. This is crucial in handling the complexities and nuances of medical diagnostics. The integration of Cross-Entropy Loss function and Reinforcement Learning from Human Feedback (RLHF) in the training process further refines the model’s accuracy and human-like understanding, addressing the high sensitivity yet lower specificity issue identified in previous models.

## Results

### Overview of Study Population

In the study involving 11,428 individuals who presented with emergency chest pain, after initially assessing 12,015 potential participants, 587 were excluded due to significant gaps in their medical records or indirect patient complaints. The study group had an average age of 64.82 years, with a broad age distribution from 15 to 101 years, highlighting a significant elderly presence, underlined by a median age of 67 years. Men constituted 65.4% of the participants.

The average Body Mass Index (BMI) for the cohort was 25.41, with a standard deviation of 3.69. Medical evaluations revealed an average systolic blood pressure of 124.28 mmHg, diastolic blood pressure of 75.35 mmHg, and heart rate of 70.38 bpm. The patient histories showed varying prevalences of conditions: 8.7% had chest pain, 15.3% experienced dyspnea or chest tightness, and 3.7% had episodes of syncope. Additionally, there were notable rates of chronic conditions, including diabetes (7.9%), hypertension (23.5%), and hyperlipidemia (17.3%). Lifestyle factors were also recorded, with 18.9% having a smoking history and 15.3% with a history of alcohol consumption. In terms of emergency severity, 2.8% of cases were classified as critical or severe, while 14.7% were urgent, and the majority, 82.5%, were less urgent. The diversity of cardiovascular conditions was evident in the primary diagnoses. (Supplementary Table S2)

The disease composition spectrum of 11,428 patients was analyzed based on the “primary diagnosis” of discharge diagnosis. The most common cardiovascular issues were NSTEMI/UA Unstable Angina (24.3%), followed by Stable Angina Pectoris (14.8%) and STMI (7.4%). Other cardiovascular diagnoses included Chronic Coronary Syndrome, Aortic Dissection, and Acute Pulmonary Embolism. Hypertensive emergencies varied in severity and risk, with a range of stages and risks documented. Arrhythmias formed a significant part of the diagnoses, with conditions like Paroxysmal and Persistent Atrial Fibrillation, Atrial Flutter, and Wolff-Parkinson-White Syndrome being prevalent. Heart failure variants were also noted, along with other cardiac conditions such as Old Myocardial Infarction and Aortic Valve Insufficiency. This detailed assessment underscores the wide spectrum of cardiovascular diseases managed in the emergency setting, reflecting the complexity and diversity of the patient population. (Supplementary Table S3)

### Performance of LLM

We assessed the normality of the LLM’s performance distribution using kurtosis, skewness, probability density curve, P-P diagram, and Q-Q diagram. (Supplementary Figure S1)

Our analysis confirmed a normal distribution without significant outliers. Regarding the diagnosis efficacy, “Tongyi Qianwen” achieved an accuracy of 61.11% (95% CI:60.84%-61.29), with a sensitivity of 91.67% (95% CI:91.37%-91.96%) and a specificity of 47.95% (95% CI:47.65%-48.25%). “Lingyi Zhihui” demonstrated an accuracy of 76.40% (95% CI:76.17%-76.63%), with a sensitivity of 90.99% (95% CI:90.67%-91.31%) and a specificity of 70.15% (95% CI:69.85%-70.44%). The human experts were asked to perform the diagnostic test based on the same content fed to LLMs. A total of 8 physicians completed this task. Human experts achieved a mean accuracy of 86.37% (95% CI:86.18%-86.55%), a sensitivity of 79.62% (95% CI:79.20%-80.04%), and a specificity of 89.26%(95% CI:89.06%-89.46%) **Table 1**.

**Table 1:** Comparison of accuracy, sensitivity, and specificity in diagnosing emergency ACS between the two large language model diagnostic groups and the human expert diagnostic group.

| Model/Groupe | Accuracy (95%CI) | Sensitivity (95%CI) | Specificity (95%CI) |
| --- | --- | --- | --- |
| TongyiQianwen (v1.0.3) | 61.11% (95% CI:60.84%-61.29%) | 91.67% (95%CI:91.37%-91.96%) | 47.95% (95%CI:47.65%-48.25%) |
| Lingyi Zhihui (v2.2.0) | 76.40% (95% CI:76.17%-76.63%) | 90.99% (95%CI:90.67%-91.31%) | 70.15% (95%CI:69.85%-70.44%) |
| Human Experts | 86.37% (95%CI:86.18%-86.55%) | 79.62% (95%CI:79.20%-80.04%) | 89.26% (95% C:89.06%-89.46%) |

This table provides a detailed comparison of the key performance metrics when diagnosing emergency ACS among the two large language model diagnostic groups and the human expert diagnostic group.

The language models “Tongyi Qianwen” and “Lingyi Zhihui” completed the task in 24.68±2.23 and 28.75±3.25 minutes, respectively. On the other hand, human physicians completed the task within a range of 65.25±10.45 minutes (**Figure 1a**).

We plotted the parameters of diagnostic performance using radar charts. The area under the curve for “Lingyi Zhihui” was 8094.76 units, which was more significant than the 5597.88 units for “Tongyi Qianwen”. However, human experts had the best overall performance, totaling 9393.36 units (**Figure 2a**). The area for “Tongyi Qianwen” performance primarily spans over the “Sensitivity” region but is relatively smaller in the “Specificity” and “Accuracy” regions. “Lingyi Zhihui” had a larger area in all three parts compared to “Tongyi Qianwen”, especially in the “Specificity” domain. The area for human experts was substantial in both the “Specificity” and “Accuracy” regions but slightly smaller in the “Sensitivity” domain.

Both “Tongyi Qianwen” and “Lingyi Zhihui” demonstrated a high level of sensitivity, indicating their capability to detect the majority of true ACS cases. However, their specificity was comparatively lower, implying the potential for misclassifying some non-ACS cases as ACS. High sensitivity is pivotal in screening tools, as they aim to identify the most genuine cases, even at the risk of producing some false positives. Ensuring the accurate detection of real diseases or abnormalities is a critical attribute of screening tools. Consequently, LLMs are well-suited as screening tools, particularly in life-threatening emergency scenarios. In such situations, the primary goal during screening is to identify as many true cases as possible, minimizing the risk of overlooking vital information. However, it’s important to acknowledge that a trade-off exists between high sensitivity and specificity, meaning that while a screening tool can capture most genuine cases, it may also generate some false positives (false alarms), which must be carefully considered. (**Figure 2 b. c**)

“Tongyi Qianwen” model achieved a true positive rate (TPR) of 91.67% and a concomitant false positive rate (FPR) of 52.05%. The model’s accuracy is 43.10%, consistent with its recall. On the other hand, “Lingyi Zhihui” shows that its TPR and recall rate are both 90.99%, but its FPR is significantly reduced to 29.85%, and its accuracy rate is 56.87%. In contrast, the human expert’s TPR was 79.62%, the FPR was reduced considerably to 10.74%, and the accuracy was 76.34%, consistent with its recall rate (**Figure 2b**).

Among the cases misdiagnosed as ACS by the “Tongyi Qianwen” test, approximately 7.34% (95% CI: 7.07%-7.60%) were eventually diagnosed as aortic dissection, and 3.45% were diagnosed as acute pulmonary embolism according the reference standard (95% CI: 3.26%-3.63%). The rest were other non-ACS diseases with chest pain manifestations. Among the cases misdiagnosed as ACS by the “Lingyi Zhihui”, the average proportion of cases that were eventually diagnosed as acute aortic dissection was approximately 9.14% (95% CI: 8.83%-9.44%). The average proportion of patients who were eventually diagnosed with acute pulmonary embolism was 2.83% (95% CI: 2.63%-3.03%).

Although human experts presented higher accuracy, there were still some cases misdiagnosed. Among the total cases misdiagnosed as ACS by human experts, acute aortic dissection accounted for 5.27% (95% CI: 4.80%-5.74%) and acute pulmonary embolism accounted for 0.96% (95% CI: 0.74%-1.18%). The discrepancies among LLMs and human experts are statistically significant. (Supplementary Figure S2)

### Advancements in Medical Large Language Models: The Performance of MedGuide-13B

After evaluating the performance of various commercially available closed-source Large Language Models in medical diagnostics, we enhanced their capabilities by improving model architecture, refining algorithms, and boosting fine-tuning and alignment techniques to increase accuracy and reduce misdiagnoses. From these comprehensive benchmarks, we distilled key insights that provided a solid foundation for the development of new language models. Consequently, we developed the MediGuide-14B model, which was derived by making precise adjustments to the Qwen-14B base model. The Qwen-14B model, known for its strong natural language understanding and problem-solving capabilities, served as an ideal starting point for the development of MediGuide-14B.

In the development process of MediGuide-14B, we first meticulously analyzed the issues encountered by existing commercial general-purpose large language models when executing medical tasks and accordingly implemented a series of targeted parameter optimizations to enhance their performance in the healthcare domain. In the initial phase, our focus was on bolstering the model’s understanding of medical terminology. This involved expanding the medical professional vocabulary database and refining the model’s processing mechanisms for these terms. During fine-tuning, we incorporated multi-turn dialogue data derived from real-world medical scenarios involving 300,000 patients, significantly enhancing the model’s professionalism and accuracy within medical contexts.

Employing supervised fine-tuning (Supervised Fine-Tuning, SFT), the fine-tuned large model showed a significant improvement in accuracy when dealing with professional medical texts compared to the original model. Subsequently, during the alignment process of the large model, we introduced reinforcement learning from human feedback technology (Reinforcement Learning from Human Feedback, RLHF) to guide the output distribution of the large model. We solicited feedback and optimization from medical experts on the model’s outputs, thereby creating a contrastive dataset imbued with human preferences, ensuring that the decision-making process of the large model not only fully leverages its reasoning capabilities but also aligns with the judgment standards of medical professionals. A reward model (Reward Model, RM) was trained on this dataset, and reinforcement learning techniques were used to conduct further fine-tuning alignment.

The aligned model (aligned model) following this process demonstrated a substantial enhancement in its generalization ability when handling actual medical data, closely adhering to the practical needs of medicine and effectively improving the accuracy of complex case analysis. Lastly, in the model’s inference process, we employed a chain-of-thought decomposition method where complex medical scenario questions posed by users were finely dissected to accurately capture key information. This helped the model better comprehend the core content and logical structure of the problem, thereby enhancing both the accuracy and relevance of its responses. After such granular decomposition, the model independently analyzed each sub-problem before synthesizing answers from all sub-problems to form a comprehensive and logically coherent final answer.

Through the above-mentioned parameter optimizations and adjustments tailored for medical tasks, MediGuide-14B has achieved a 44% improvement in capability over its predecessors. (Supplementary Figure S3)

To assess the efficacy of large language models in diagnosing cardiovascular diseases, we constructed the CVIDB test set, comprising 1,233 single-choice questions and 203 multiple-choice questions. This standardized and high-quality test set provides detailed explanations for each question, offering insights into the reasoning behind the correct answers and enhancing learning and understanding of complex topics. The test set covers various subtypes, developmental stages, and related complications and comorbidities of cardiovascular diseases, effectively testing the depth and breadth of large language models’ understanding of the field. We have made this test set publicly available on GitHub for researchers and developers to download free of charge. The access link is: https://github.com/mengxiangbin123/CVIDB.git

After completing the training and development of the MediGuide large model, we conducted a series of standardized assessments, including several important medical benchmark tests. These tests are^55,62,63^: USMLE, a repository of simulated questions for the United States Medical Licensing Examination; MedMCQA, a large-scale medical multiple-choice question dataset covering various disciplines, derived from medical entrance exams in India; CMC, a large-scale multitask knowledge assessment benchmark focusing on Chinese medical knowledge; and MCMLE, a simulation of the Chinese medical qualification exam; along with the cardiovascular disease-specific benchmark test set, CVIDB. These resources aim to comprehensively evaluate the performance and generalization ability of large language models in medical knowledge and clinical decision-making skills. We compared the performance of MediGuide-14B (V5.0) with other leading models in the industry, including ChatGPT-4, ChatGPT-3.5 Turbo, Tongyi Qianwen (v1.0.3), Lingyi Zhihui (v2.2.0), LLaMA 2-14B, and Qianwen-14B -Base.

Focusing on the United States Medical Licensing Examination (USMLE), ChatGPT-4 demonstrated superior performance with a score of 80.28%, closely followed by MediGuide-14B at 78.63%, while LLaMA 2-13B trailed significantly with only 35.04%. For the MedMCQA, which consists of multiple-choice questions from Indian medical entrance exams, ChatGPT-4 again led with a score of 72.51%, although here, the performance differences among the newer models were relatively narrower. In contrast, models like ChatGPT-3.5 Turbo and Qianwen-14B -Base showed relatively lower scores, 56.25% and 42.86%, respectively. The Composite Medical Content (CMC) dataset, which assesses the models’ understanding of medical knowledge specifically in the Chinese context, saw MediGuide-14B performing the best with a score of 77.56%. ChatGPT-4 and Lingyi Zhihui also showed strong results with scores above 73%. Performance on the Medical Chinese Medical Licensing Examination (MCMLE) again highlighted the effectiveness of ChatGPT-4 and MediGuide-14B, which scored 74.58% and 75.41%, respectively, demonstrating their robustness in handling questions related to the Chinese Medical Licensing Examination. Lower-tier models, such as LLaMA 2-13B, had notably weaker performance, indicating possible challenges in their language-specific medical knowledge. Lastly, in the Cardiovascular Disease Intelligence Diagnostic Benchmark (CVIDB), MediGuide-14B exhibited exceptional capability, scoring the highest at 80.85%, showing its potential utility in applications focused on cardiovascular health. ChatGPT-4 remained consistent across all benchmarks with scores generally above 75%, reinforcing its overall reliability in medical domain question answering. (Supplementary Table S4).

MediGuide-14B underwent a thorough evaluation process like that of ‘Tongyi Qianwen’ and ‘Lingyi Zhihui.’ It was tested using 1000 test units, each consisting of 100 distinct real-world cases sourced from actual medical scenarios. This rigorous testing framework provided a comprehensive assessment of MediGuide-14B’s performance in real-life conditions. The model achieved an impressive accuracy rate of 84.52%. It demonstrated high sensitivity in correctly identifying positive results and commendable specificity in correctly identifying negative results (Supplementary Figure S4).

### Extended recommendations by LLMs

For the reference standards, we invited a panel of four distinguished cardiovascular specialists, each with over twenty years of clinical experience. To further evaluate the possibility of LLM’s role in emergency Chest Pain Triage, we asked them to arbitrarily evaluate the treatment recommendations generated from the prompts. The evaluation was based on established guidelines for diagnosing and treating chest pain and full access to an array of essential patient data: from electrocardiograms (ECGs) and cardiac enzyme tests to echocardiograms, NT-proBNP evaluations, and when indicated, results from coronary angiography.

As illustrated in Supplementary Figure S5, 3.32% (95% CI:3.22%-3.41%) of the recommendations generated by “Tongyi Qianwen” were deemed unsuitable. This resulted in significant omissions of critical content that could potentially endanger patients. However, 12.88% (95% CI:12.71%-13.04%) of the recommendations were considered reasonable, albeit incomplete, with no direct harm to patients. The remaining 83.81% (95% CI:83.64%-83.98%) of recommendations were classified as comprehensive and appropriate.

Regarding the “Lingyi Zhihui” model, 3.40% (95% CI:3.30%-3.50%) of recommendations were deemed inappropriate with inherent risks. 43.44% (95% CI:43.18%-43.69%) were considered reasonable but not exhaustive, devoid of direct patient harm. Meanwhile, 53.16% (95% CI:52.91%-53.41%) of recommendations were thoroughly comprehensive and relevant.

The diagnostic suggestions from human experts were deemed that 2.48% (95% CI:0.28%-4.68%) were inappropriate and could potentially compromise timely patient treatment. Another 12.96% (95% CI:7.69%-18.23%) were considered reasonable but not comprehensive, while a substantial 84.56% (95% CI:74.45%-94.67%) were acknowledged as fully comprehensive and appropriate. The performance assessment of the MediGuide-14B group in terms of treatment recommendations reveals a nuanced picture. A small fraction, specifically 2.90% (95% CI:1.86%-3.94%), of the recommendations were categorized as unreasonable and risky, highlighting areas where caution is necessary. On the other hand, 13.52% (95% CI:11.38%-15.64%) of the recommendations were deemed reasonable, albeit incomplete, suggesting a foundation of sound medical guidance that could benefit from further elaboration or additional information. The majority, 83.58% (95% CI:81.28%-85.98%) of the recommendations from the MediGuide-14B group stood out as both reasonable and comprehensive, indicating a high level of proficiency in providing well-rounded and thorough treatment advice. (Supplementary Figure S5)

### The impact of employment status of patients on the ACS Diagnostic Accuracy of LLMs

Next, we sought to evaluate the impact of employment status of patients on LLM diagnostic accuracy. We hypothesized that employment status may be linked to inherent characteristics that could influence the information extracted from a patient’s chief complaint. Given that individuals covered by the Urban and Rural Resident Basic Medical Insurance (URRBMI) are typically unemployed or self-employed, while those covered by the Urban Employee Basic Medical Insurance (UEBMI) are typically employed by institutions, we leveraged health insurance data extracted from medical records to infer the employment status of patients^64,65^. In the case of patients under the URRBMI insurance plan, “Tongyi Qianwen” exhibited a diagnostic accuracy of 58.56%, while it demonstrated a slightly higher accuracy of 60.74% among patients under the UEBMI plan (P<0.05). Interestingly, the diagnostic accuracy of “Lingyi Zhihui” was also affected by the employment status of the patients: an accuracy rate of 77.62% under URRBMI and 74.75% under UEBMI (P<0.05).

In the evaluation of MediGuide-14B’s performance across different insurance types, the group demonstrated notable results in the realm of Supplementary Diagnosis accuracy. For cases covered under the URRBMI, the MediGuide-14B achieved a mean accuracy of 83.65%, under the UEBMI category, the mean accuracy recorded was slightly higher, at 85.04% (P=0.046). (Supplementary Figure S6)

### The impact of patient’s history on LLMs diagnosis efficacy

Medical history is crucial in diagnosis, offering insights into a patient’s past health and disease risk factors. While human doctors can interpret this data based on experience, LLMs face a challenge in doing so effectively. We sought to analyze how medical history affects LLMs’ diagnosis performance.

In the absence of medical history, “Tongyi Qianwen” initially demonstrated a mean accuracy of 72.66% (95% CI: 72.43%-72.90%), a sensitivity of 81.65% (95% CI: 81.25%-82.06%), and a specificity of 68.81% (95% CI: 68.53%-69.09%). Upon the inclusion of past medical histories, the accuracy decreased to 61.110% (95% CI: 60.84%-61.29%). However, sensitivity increased to 91.67% (95% CI: 91.37%-91.96%), while specificity decreased to 47.95% (95% CI: 47.65%-48.25%). Additional details can be found in Figure 3. In the case of “Lingyi Zhihui,” without medical history, the diagnostic accuracy was 74.17% (95% CI: 73.94%-74.39%), sensitivity stood at 89.06% (95% CI: 88.73%-89.40%), and specificity at 67.78% (95% CI: 67.49%-68.07%) as depicted in Figure 3. Subsequently, with the incorporation of a more comprehensive dataset, “Lingyi Zhihui” achieved an accuracy of 76.40% (95% CI: 76.17%-76.63%), sensitivity of 90.99% (95% CI: 90.68%-91.31%), and specificity of 70.15% (95% CI: 69.85%-70.44%).

Regarding the “Tongyi Qianwen” model, the initial treatment suggestions in the absence of medical history were deemed inappropriate and potentially harmful in 5.80% of instances (95% CI: 5.68%-5.93%). Approximately 51.33% (95% CI: 51.11%-51.55%) were considered reasonable but incomplete, while 42.87% (95% CI: 42.64%-43.09%) were assessed as both comprehensive and suitable. In subsequent recommendations when medical history was provided, the figures shifted to 3.32% (95% CI: 3.22%-3.41%) being inappropriate, 12.88% (95% CI: 12.71%-13.04%) being reasonable but partial, and 83.81% (95% CI: 83.64%-83.98%) being thorough and appropriate (Fig. 3). For the “Lingyi Zhihui” model, the recommendations based on prompts without medical history were categorized as inappropriate in 6.50% (95% CI: 6.37%-6.63%) cases, reasonable but lacking in 28.45% (95% CI: 28.21%-28.69%), and both comprehensive and fitting in 65.05% (95% CI: 64.80%-65.31%). When medical history was provided, “Lingyi Zhihui” recommendations shifted to 3.40% (95% CI: 3.30%-3.50%) being inappropriate, 43.44% (95% CI: 43.19%-43.68%) being reasonable but not exhaustive, and 53.16% (95% CI: 52.91%-53.41%) being comprehensive and relevant. (Figure 3)

The removal of past medical history significantly impacted “Tongyi Qianwen,” notably increasing specificity by 20.86% and accuracy by 11.55%, while sensitivity declined by 10.02%. Conversely, “Lingyi Zhihui” exhibited relatively minor changes, with specificity decreasing by 2.37%, accuracy by 2.23%, and sensitivity by 1.93% (Supplementary Figure S7). Following the omission of medical history, “Tongyi Qianwen” reduced the rate of inappropriate treatment recommendations from 5.80% to 3.32%, while comprehensive and appropriate recommendations surged from 42.87% to 83.81%. In contrast, “Lingyi Zhihui” saw a decline in inappropriate recommendations from 6.50% to 3.40%, but comprehensive and appropriate recommendations decreased from 65.05% to 53.16%. This indicates that both models decreased the frequency of inappropriate recommendations when excluding medical history, with “Tongyi Qianwen” notably enhancing comprehensive and appropriate suggestions while “Lingyi Zhihui” experienced a decline.

In assessing the impact of removing past medical history on MediGuide-14B’s model metrics, a series of changes were observed. The accuracy of the model experienced a decrease of 2.90%. Additionally, there was a 4.26% reduction in sensitivity, indicating a diminished capacity of the model to correctly identify positive cases. Finally, the model’s specificity also decreased by 1.58%, reflecting a slight reduction in its ability to accurately identify negative cases. (Supplementary Figure S7)

## Discussion

As large language models (LLMs) continue to advance and find widespread application, they have demonstrated transformative potential in medical tasks. However, evaluating the capabilities of large language models is a complex and challenging scientific issue. Currently, the assessment of these models’ medical knowledge and logical reasoning abilities primarily relies on standardized tests. Yet, real-world medical tasks often exceed the scope of structured tasks, presenting a high level of complexity and uncertainty. There is a global lack of systematic evaluation of large language models’ effectiveness in actual medical settings. Moreover, the pre-training data for the world’s major language models is predominantly in English. For instance, the recently released Llama3 has about 5% of its corpus in non-English languages; ChatGPT-3.5 has approximately 0.09905% of its pre-training data in Chinese. Even models intended primarily for Chinese contexts, such as Tongyi Qianwen, Wenxin Yiyen, and Baichuan, have only 15-30% of their datasets in Chinese. Considering the structural, grammatical, and usage differences between non-segmented and segmented texts, the composition of different language families in LLMs’ pre-training datasets might affect their performance in various linguistic environments, which is a scientific question worthy of in-depth discussion.

This study aims to fill these gaps, focusing on the specific applications of large language models in emergency triage or consultation scenarios. This study compares the diagnostic performance of AI-driven models and human expertise in triaging emergency chest pain cases. While previous research has primarily focused on English-based ChatGPT models, this study is pioneering in evaluating two LLMs designed for “non-segmented text” environments.

Traditional machine learning systems (MLS) use specific structured data from the Electronic Emergency Triage System (EETS) to enhance the identification of critically ill patients. These MLS employ a predictive model primarily using the CatBoost Python package and provide real-time explanations via the SHAP method to help medical staff understand why certain patients may require immediate treatment. However, these systems have limitations, such as potential overfitting issues, a lack of effective capture of complex nonlinear relationships, and challenges in processing unstructured data. While models built on traditional machine learning or deep learning perform well on specific datasets, they generally lack generalizability and often show reduced predictive power when patient populations and samples are changed. This is why currently, there are no truly integrated predictive models in medical systems worldwide, and a significant gap exists between academic research on predictive models and their clinical applications^25^. Models like GPT-4 and similar large language models, with their strong capabilities in understanding and generating natural language, logical reasoning, and knowledge storage, show significant advantages in handling various data types, including unstructured, multimodal, and dynamic data. It’s worth noting that “Tongyi Qianwen (LLM)” and “Lingyi Zhihui (LLM)” exhibited high sensitivity but lower specificity, particularly when compared to human experts. This raises concerns about potential overdiagnosis by the AI models, which could result in unnecessary tests and treatments. However, high sensitivity is crucial for screening tools, as they aim to capture most of the true cases, even if it leads to some false positives. The high sensitivity of the AI models suggests their suitability as initial diagnostic tools to ensure potential positive cases are not missed. While both LLMs demonstrated the ability to provide relevant medical advice, there were notable differences in the depth and validity of their recommendations. This underscores the need to optimize LLMs for medical scenarios before deployment in healthcare settings. It is essential to utilize more realistic medical data during training and fine-tune the models to align with the nuances of medical treatment itself ^66,67^.

This study found that LLMs struggled to significantly improve diagnostic accuracy and treatment recommendations when incorporating patients’ medical histories. This could be attributed to two key factors: Current LLMs rely on computational power and probabilistic calculations rather than a deep understanding of disease mechanisms. Second, there’s a need for more advanced algorithms that can better extract relevant clinical information while filtering out noise from historical data. The removal of past medical history significantly impacted “Tongyi Qianwen,” leading to a substantial increase in specificity and accuracy while decreasing sensitivity. Meanwhile, “Lingyi Zhihui” exhibited minor changes in diagnostic metrics. Additionally, both models altered the frequency of inappropriate treatment recommendations when medical history was omitted, with “Tongyi Qianwen” improving its comprehensive and appropriate suggestions, while “Lingyi Zhihui” declined. When utilizing LLMs for diagnostic support, healthcare practitioners should acknowledge variations in how these models handle intricate and diverse data^68^. Factors such as model comprehensiveness, accuracy, and potential sources of interference should be considered. Clinical judgment, rooted in experience, should guide decision-making. Continuous monitoring and performance optimization are crucial. LLMs offer promise as diagnostic aids, but healthcare professionals must weigh multiple factors to ensure the delivery of precise and thorough diagnostic recommendations to patients.

Our study shifted focus to MediGuide-14B, our proprietary open-source model. This model has been specifically fine-tuned for medical applications, providing us with an opportunity to scrutinize its real-world efficacy. In our preceding analyses, we observed notable variations in sensitivity and specificity across different LLMs, underscoring the need for a detailed examination of each model’s strengths and weaknesses. MediGuide-14B stands out as a large language model dedicated to the medical sector, boasting an advanced integration of domain-specific datasets and bespoke training approaches to optimize its diagnostic capabilities.

Conducting an exhaustive evaluation of MediGuide-14B’s performance is pivotal not only for gauging the broader applicability of LLMs in healthcare but also for charting the course for their future development and potential areas of application. By juxtaposing MediGuide-14B against other leading models in the field, we aim to deliver a nuanced appraisal of the model’s accuracy, efficiency, and reliability in medical diagnostics. This comparative analysis is intended to furnish diverse insights and formative experiences, contributing significantly to the ongoing discourse on the role and impact of large language models in healthcare research.

Our study illustrates the marked variability in the performance of different large language models when processing real-world medical scenario information. The objective of our research is not solely to compare and rank these models but to emphasize the adaptability and potential of LLMs in the medical field. This realization underscores the necessity of careful and precise benchmarking tailored for medical AI applications. We have also discovered that specific fine-tuning and alignment of LLMs significantly enhance their ability to perform specialized tasks within the medical domain, even on smaller-scale models. This finding is particularly significant for vertical sectors like healthcare, as it suggests the feasibility of training and deploying efficient medical LLMs at a lower cost. Such advancements allow us to apply cutting-edge AI technology more effectively in clinical settings, thereby improving the quality and efficiency of healthcare services.

LLMs have the potential to reshape certain aspects of healthcare, particularly in the context of rapid pre-hospital Chest Pain Triage. The integration of these models has the potential to streamline triage procedures, facilitating timely interventions even before a patient arrives at the hospital. This not only enhances the effectiveness and scope of diagnostic and therapeutic interventions but also promises to improve the efficiency of medical infrastructure, reduce patient waiting times, alleviate the burden on emergency medical personnel, and ultimately alleviate the financial strain on patients and healthcare systems^69–71^. In conclusion, the diagnostic capabilities demonstrated by LLMs, as evidenced in this study, underscore their significance in advancing the field of rapid triage. It is reasonable to anticipate that soon, these models will play a pivotal role in enhancing healthcare delivery, ultimately benefiting both patients and healthcare systems.

In an era increasingly dominated by AI, medical practitioners, particularly the younger generation, will inevitably encounter an expanding array of medical AI entities. How they utilize AI, discern which AI tools best assist them, and identify the specific functions where AI can provide support, necessitates robust benchmarking efforts. Such benchmarks offer crucial guidance to healthcare professionals in navigating the AI landscape. Our study aims to initiate this journey in the field, laying a foundational step that we believe will serve as a vital reference for future researchers. This work is poised to propel the further advancement and application of LLMs in healthcare, ultimately aiding medical professionals in harnessing AI’s full potential for improved patient care and healthcare delivery.

### Limitations

The reliance on retrospective data may introduce inherent biases, potentially impacting the generalizability of the results. Additionally, LLMs’ diagnostic performance in ACS triaging scenarios might not reflect their capabilities in other medical conditions. The specificity challenges highlighted in the study emphasize the need for broader and more diverse training datasets. Furthermore, the comparative analysis between AI models and human experts, though illuminating, is based on a restricted set of parameters, potentially overlooking nuanced aspects of clinical decision-making. The study underscores the necessity of comprehensive, prospective research to validate the findings and address these limitations.

## Data Availability

All data produced in the present study are available upon reasonable request to the authors

## Declarations

This manuscript was edited by LLM “Tongyi Qianwen” for its English language, but human authors read and made the final version.

## Author Contributions

The study was primarily designed by Yi-Da Tang, Xiangbin Meng, Xiangyu Yan, and Jun Gao.

## Individual Contributions

- Yi-Da Tang: Led the study design, contributed cardiovascular expertise, involved in data collection and analysis, reviewed and approved the final manuscript.
- Xiangbin Meng: Contributed cardiovascular expertise, involved in data collection and analysis, reviewed and approved the final manuscript.
- Xiangyu Yan: Provided domain-specific knowledge, involved in data collection, analysis, and interpretation, reviewed and approved the final manuscript.
- Jun Gao: Contributed cardiovascular expertise, involved in data collection and analysis, reviewed and approved the final manuscript.
- Jingjia Wang, Xuliang Wang, Yuan-geng-shuo Wang, Wenyao Wang, Chunli Shao: Contributed cardiovascular expertise, were involved in data collection and analysis, reviewed and approved the final manuscript.
- Junhong Wang, Yaodong Yang, Muhan Zhang, Xiaojuan Cui, Jing Chen, Kuo Zhang, Da Liu, Jia-ming Ji, Zifeng Qiu, Muzi Li: Provided domain-specific knowledge in data collection, analysis, and interpretation, reviewed and approved the final manuscript.

## Data Verification

Yi-Da Tang、Xiangbin Meng and Jun Gao directly accessed and verified the underlying data reported in the manuscript, ensuring its integrity.

## Full Access and Responsibility

All authors confirm that they had full access to all the data in the study and accept the responsibility for the decision to submit for publication.

## Collaboration and Coauthorship

The authors acknowledge and appreciate the collaboration and coauthorship of colleagues in the locations where the research was conducted, reflecting the benefits of diversity in authorship in terms of background, career-stage, gender, geography, and race.

## Acknowledgments and Personal Communications

All cited individuals in acknowledgments or personal communications have provided their written consent.

## Author Statement Form

All authors have signed the author statement form, which will be uploaded with the submission.

## Acknowledgments

This study was funded by the National Key R&D Program of China (2020YFC2004705), National Natural Science Foundation of China (81825003, 91957123, 82270376), CAMS Innovation Fund for Medical Sciences (2022-I2M-C&T-B-119, 2021-I2M-5-003), Beijing Nova Program (Z201100006820002) from Beijing Municipal Science & Technology Commission, and CSC Special Fund for Clinical Research (CSCF2021A04).

---

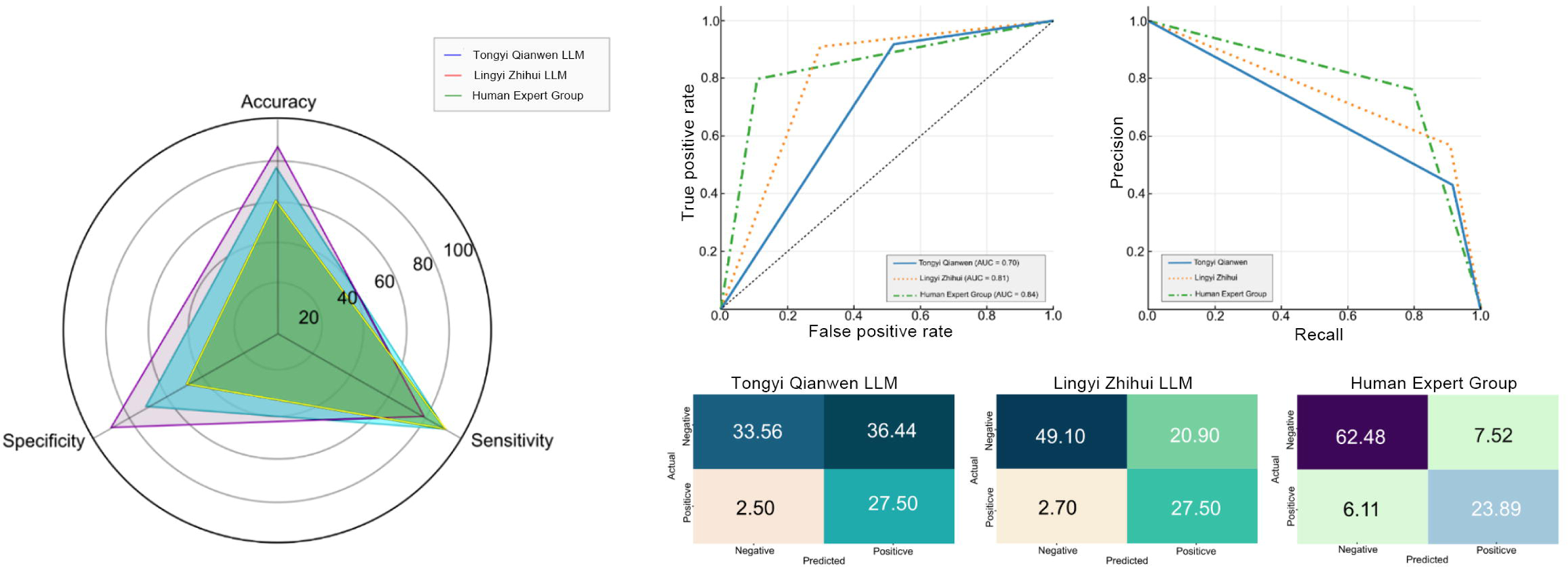

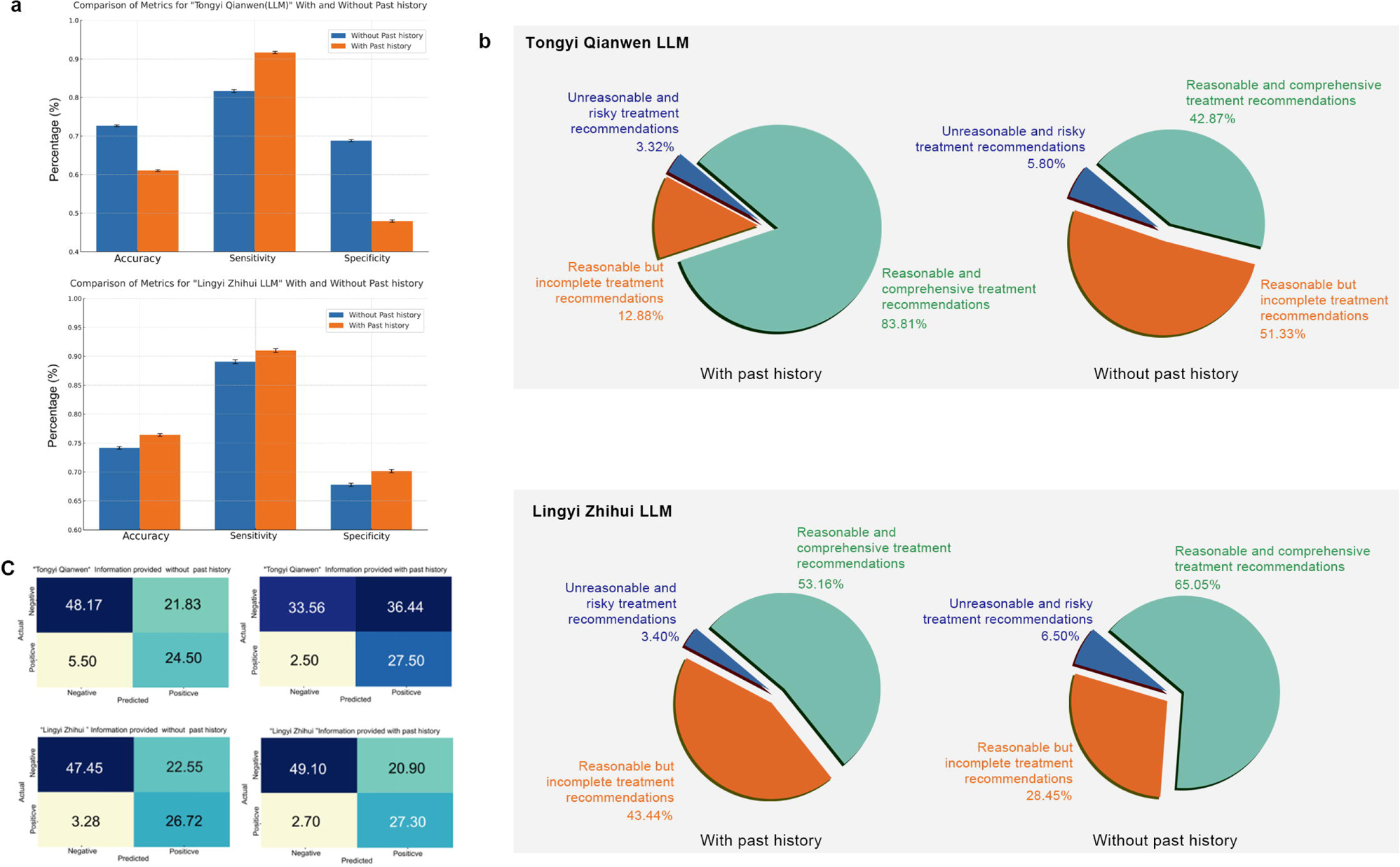

## References

1. Ayers JW, Zhu Z, Poliak A, et al. Evaluating Artificial Intelligence Responses to Public Health Questions. JAMA Netw Open 2023; 6(6): e2317517.

2. Kanjee Z, Crowe B, Rodman A. Accuracy of a Generative Artificial Intelligence Model in a Complex Diagnostic Challenge. Jama 2023; 330(1): 78–80.

3. Minssen T, Vayena E, Cohen IG. The Challenges for Regulating Medical Use of ChatGPT and Other Large Language Models. Jama 2023.

4. Will ChatGPT transform healthcare? Nat Med 2023; 29(3): 505–6.

5. Gilbert S, Harvey H, Melvin T, Vollebregt E, Wicks P. Large language model AI chatbots require approval as medical devices. Nat Med 2023.

6. Thirunavukarasu AJ, Ting DSJ, Elangovan K, Gutierrez L, Tan TF, Ting DSW. Large language models in medicine. Nat Med 2023.

7. Singhal K, Azizi S, Tu T, et al. Large language models encode clinical knowledge. Nature 2023.

8. Howard A, Hope W, Gerada A. ChatGPT and antimicrobial advice: the end of the consulting infection doctor? Lancet Infect Dis 2023; 23(4): 405–6.

9. Arora A, Arora A. The promise of large language models in health care. The Lancet 2023; 401(10377): 641.

10. Jiang LY, Liu XC, Nejatian NP, et al. Health system-scale language models are all-purpose prediction engines. Nature 2023; 619(7969): 357–62.

11. Thapa S, Adhikari S. ChatGPT, Bard, and Large Language Models for Biomedical Research: Opportunities and Pitfalls. Ann Biomed Eng 2023.

12. Ouyang L, Wu J, Jiang X, et al. Training language models to follow instructions with human feedback. arXiv pre-print server 2022.

13. Wayne, Zhou K, Li J, et al. A Survey of Large Language Models. arXiv pre-print server 2023.

14. Sharma P, Parasa S. ChatGPT and large language models in gastroenterology. Nat Rev Gastroenterol Hepatol 2023.

15. Li R, Kumar A, Chen JH. How Chatbots and Large Language Model Artificial Intelligence Systems Will Reshape Modern Medicine: Fountain of Creativity or Pandora’s Box? JAMA Intern Med 2023; 183(6): 596–7.

16. Wei J, Wang X, Schuurmans D, et al. Chain-of-thought prompting elicits reasoning in large language models. Advances in Neural Information Processing Systems 2022; 35: 24824–37.

17. Miller K, Gunn E, Cochran A, et al. Use of Large Language Models and Artificial Intelligence Tools in Works Submitted to Journal of Clinical Oncology. Journal of clinical oncology : official journal of the American Society of Clinical Oncology 2023; 41(19): 3480–1.

18. Ayers JW, Poliak A, Dredze M, et al. Comparing Physician and Artificial Intelligence Chatbot Responses to Patient Questions Posted to a Public Social Media Forum. JAMA Intern Med 2023; 183(6): 589–96.

19. Wei J, Tay Y, Bommasani R, et al. Emergent Abilities of Large Language Models. arXiv pre-print server 2022.

20. Azizi Z, Alipour P, Gomez S, et al. Evaluating Recommendations About Atrial Fibrillation for Patients and Clinicians Obtained From Chat-Based Artificial Intelligence Algorithms. Circulation: Arrhythmia and Electrophysiology 2023: e012015.

21. Yang X, Chen A, PourNejatian N, et al. A large language model for electronic health records. NPJ Digit Med 2022; 5(1): 194.

22. Sheikh K, Ghaffar A. PRIMASYS: a health policy and systems research approach for the assessment of country primary health care systems. Health Research Policy and Systems 2021; 19(1): 31.

23. Mehmood A, Rowther AA, Kobusingye O, Hyder AA. Assessment of pre-hospital emergency medical services in low-income settings using a health systems approach. International Journal of Emergency Medicine 2018; 11(1): 53.

24. Gizaw Z, Astale T, Kassie GM. What improves access to primary healthcare services in rural communities? A systematic review. BMC Primary Care 2022; 23(1): 313.

25. Markowetz F. All models are wrong and yours are useless: making clinical prediction models impactful for patients. npj Precision Oncology 2024; 8(1): 54.

26. Yeo YH, Samaan JS, Ng WH, et al. GPT-4 outperforms ChatGPT in answering non-English questions related to cirrhosis. 2023.

27. Fang C, Ling J, Zhou J, et al. How does ChatGPT4 preform on Non-English National Medical Licensing Examination? An Evaluation in Chinese Language. 2023.

28. Bijani M, Abedi S, Karimi S, Tehranineshat B. Major challenges and barriers in clinical decision-making as perceived by emergency medical services personnel: a qualitative content analysis. BMC Emergency Medicine 2021; 21(1): 11.

29. Becker TK, Gausche-Hill M, Aswegan AL, et al. Ethical challenges in Emergency Medical Services: controversies and recommendations. Prehosp Disaster Med 2013; 28(5): 488–97.

30. Pines JM, Mullins PM, Cooper JK, Feng LB, Roth KE. National trends in emergency department use, care patterns, and quality of care of older adults in the United States. Journal of the American Geriatrics Society 2013; 61(1): 12–7.

31. Vainieri M, Panero C, Coletta L. Waiting times in emergency departments: a resource allocation or an efficiency issue? BMC Health Serv Res 2020; 20(1): 549.

32. Neprash HT, Everhart A, McAlpine D, Smith LB, Sheridan B, Cross DA. Measuring Primary Care Exam Length Using Electronic Health Record Data. Med Care 2021; 59(1): 62–6.

33. Zhiting G, Jingfen J, Shuihong C, Minfei Y, Yuwei W, Sa W. Reliability and validity of the four-level Chinese emergency triage scale in mainland China: A multicenter assessment. International Journal of Nursing Studies 2020; 101: 103447.

34. Tanabe P, Gimbel R, Yarnold PR, Kyriacou DN, Adams JG. Reliability and validity of scores on The Emergency Severity Index version 3. Academic emergency medicine 2004; 11(1): 59–65.

35. Taboulet P, Maillard-Acker C, Ranchon G, et al. Triage des patients à l’accueil d’une structure d’urgences. Présentation de l’échelle de tri élaborée par la Société française de médecine d’urgence: la FRench Emergency Nurses Classification in Hospital (FRENCH). Annales françaises de médecine d’urgence 2019; 9(1): 51–9.

36. Kachalia A, Gandhi TK, Puopolo AL, et al. Missed and delayed diagnoses in the emergency department: a study of closed malpractice claims from 4 liability insurers. Annals of emergency medicine 2007; 49(2): 196–205.

37. Hussain F, Cooper A, Carson-Stevens A, et al. Diagnostic error in the emergency department: learning from national patient safety incident report analysis. BMC Emerg Med 2019; 19(1): 77.

38. Gulati M, Levy PD, Mukherjee D, et al. 2021 AHA/ACC/ASE/CHEST/SAEM/SCCT/SCMR Guideline for the Evaluation and Diagnosis of Chest Pain: Executive Summary: A Report of the American College of Cardiology/American Heart Association Joint Committee on Clinical Practice Guidelines. Journal of the American College of Cardiology 2021; 78(22): 2218–61.

39. Amsterdam EA, Wenger NK, Brindis RG, et al. 2014 AHA/ACC guideline for the management of patients with non–ST-elevation acute coronary syndromes: a report of the American College of Cardiology/American Heart Association Task Force on Practice Guidelines. Journal of the American College of Cardiology 2014; 64(24): e139–e228.

40. Lawton JS, Tamis-Holland JE, Bangalore S, et al. 2021 ACC/AHA/SCAI guideline for coronary artery revascularization: executive summary: a report of the American College of Cardiology/American Heart Association Joint Committee on Clinical Practice Guidelines. Circulation 2022; 145(3): e4–e17.

41. Li J, Li X, Wang Q, et al. ST-segment elevation myocardial infarction in China from 2001 to 2011 (the China PEACE-Retrospective Acute Myocardial Infarction Study): a retrospective analysis of hospital data. The Lancet 2015; 385(9966): 441–51.

42. Komorowski M, Del Pilar Arias López M, Chang AC. How could ChatGPT impact my practice as an intensivist? An overview of potential applications, risks and limitations. Intensive Care Med 2023; 49(7): 844–7.

43. Madden MG, McNicholas BA, Laffey JG. Assessing the usefulness of a large language model to query and summarize unstructured medical notes in intensive care. Intensive Care Med 2023.

44. Patel SB, Lam K. ChatGPT: the future of discharge summaries? Lancet Digit Health 2023; 5(3): e107–e8.

45. Ali SR, Dobbs TD, Hutchings HA, Whitaker IS. Using ChatGPT to write patient clinic letters. Lancet Digit Health 2023; 5(4): e179–e81.

46. van Heerden AC, Pozuelo JR, Kohrt BA. Global Mental Health Services and the Impact of Artificial Intelligence-Powered Large Language Models. JAMA psychiatry 2023; 80(7): 662–4.

47. Kwok KO, Wei WI, Tsoi MTF, et al. How can we transform travel medicine by leveraging on AI-powered search engines? Journal of Travel Medicine 2023; 30(4).

48. Liu F, Panagiotakos D. Real-world data: a brief review of the methods, applications, challenges and opportunities. BMC Med Res Methodol 2022; 22(1): 287.

49. Li S. Exploring the clinical capabilities and limitations of ChatGPT: a cautionary tale for medical applications. Int J Surg 2023.

50. Wang G, Yang G, Du Z, Fan L, Li X. ClinicalGPT: Large Language Models Finetuned with Diverse Medical Data and Comprehensive Evaluation. arXiv pre-print server 2023.

51. Shea YF, Lee CMY, Ip WCT, Luk DWA, Wong SSW. Use of GPT-4 to Analyze Medical Records of Patients With Extensive Investigations and Delayed Diagnosis. JAMA Netw Open 2023; 6(8): e2325000.

52. Zhao WX, Zhou K, Li J, et al. A survey of large language models. arXiv preprint arXiv:230318223 2023.

53. Sun Y, Wang S, Feng S, et al. Ernie 3.0: Large-scale knowledge enhanced pre-training for language understanding and generation. arXiv preprint arXiv:210702137 2021.

54. Wang S, Sun Y, Xiang Y, et al. Ernie 3.0 titan: Exploring larger-scale knowledge enhanced pre-training for language understanding and generation. arXiv preprint arXiv:211212731 2021.

55. Singhal K, Azizi S, Tu T, et al. Large language models encode clinical knowledge. Nature 2023; 620(7972): 172–80.

56. Rudolph J, Tan S, Tan S. War of the chatbots: Bard, Bing Chat, ChatGPT, Ernie and beyond. The new AI gold rush and its impact on higher education. Journal of Applied Learning and Teaching 2023; 6(1).

57. Chien AA, Lin L, Nguyen H, Rao V, Sharma T, Wijayawardana R. Reducing the Carbon Impact of Generative AI Inference (today and in 2035). Proceedings of the 2nd Workshop on Sustainable Computer Systems; 2023; 2023. p. 1–7.

58. Peng C, Yang X, Chen A, et al. A study of generative large language model for medical research and healthcare. npj Digital Medicine 2023; 6(1): 210.

59. Li X, Fan Y, Cheng S. AIGC In China: Current Developments And Future Outlook. arXiv preprint arXiv:230808451 2023.

60. Collet JP, Thiele H, Barbato E, et al. 2020 ESC Guidelines for the management of acute coronary syndromes in patients presenting without persistent ST-segment elevation. Eur Heart J 2021; 42(14): 1289–367.

61. Association EMBotCM, Association CPBotCHIEP. Expert consensus on emergency diagnosis and treatment of acute chest pain. Chinese Journal of Emergency Medicine 2019; 28(4): 413–20.

62. Yang A, Xiao B, Wang B, et al. Baichuan 2: Open large-scale language models. arXiv preprint arXiv:230910305 2023.

63. Mbakwe AB, Lourentzou I, Celi LA, Mechanic OJ, Dagan A. ChatGPT passing USMLE shines a spotlight on the flaws of medical education. Public Library of Science San Francisco, CA USA; 2023. p. e0000205.

64. Yu H. Universal health insurance coverage for 1.3 billion people: What accounts for China’s success? Health policy 2015; 119(9): 1145–52.

65. He W. Does the immediate reimbursement of medical insurance reduce the socioeconomic inequality in health among the floating population? Evidence from China. International Journal for Equity in Health 2023; 22(1): 1–14.

66. Su P, Vijay-Shanker K. Investigation of improving the pre-training and fine-tuning of BERT model for biomedical relation extraction. BMC Bioinformatics 2022; 23(1): 120.

67. Wu C, Zhang X, Zhang Y, Wang Y, Xie W. PMC-LLaMA: Further Finetuning LLaMA on Medical Papers. arXiv pre-print server 2023.

68. Ferryman K, Mackintosh M, Ghassemi M. Considering Biased Data as Informative Artifacts in AI-Assisted Health Care. N Engl J Med 2023; 389(9): 833–8.

69. The, Lancet. AI in medicine: creating a safe and equitable future. Lancet 2023; 402(10401): 503.

70. Han T, Lisa, Papaioannou J-M, et al. MedAlpaca -- An Open-Source Collection of Medical Conversational AI Models and Training Data. arXiv pre-print server 2023.

71. Yunxiang L, Zihan L, Kai Z, Ruilong D, You Z. ChatDoctor: A Medical Chat Model Fine-tuned on LLaMA Model using Medical Domain Knowledge. arXiv pre-print server 2023.

